# Association between upper and lower respiratory disease among patients with primary ciliary dyskinesia: an international study

**DOI:** 10.1101/2023.09.21.23295895

**Authors:** Yin Ting Lam, Jean-François Papon, Mihaela Alexandru, Andreas Anagiotos, Miguel Armengot, Mieke Boon, Andrea Burgess, Doriane Calmes, Suzanne Crowley, Sinan Ahmed. D. Dheyauldeen, Nagehan Emiralioglu, Ela Erdem Eralp, Christine van Gogh, Yasemin Gokdemir, Eric G. Haarman, Amanda Harris, Isolde Hayn, Hasnaa Ismail-Koch, Bülent Karadag, Céline Kempeneers, Elisabeth Kieninger, Sookyung Kim, Natalie Lorent, Ugur Ozcelik, Charlotte Pioch, Johanna Raidt, Ana Reula, Jobst Roehmel, Synne Sperstad Kennelly, Panayiotis Yiallouros, the EPIC-PCD team, Myrofora Goutaki

## Abstract

**Introduction:** Nearly all patients with primary ciliary dyskinesia (PCD) report ear-nose-throat (ENT) symptoms. However, scarce evidence exists about how ENT symptoms relate to pulmonary disease in PCD. We explored possible associations between upper and lower respiratory disease among patients with PCD in a multicentre study.

**Methods:** We included patients from the ENT Prospective International Cohort (EPIC-PCD). We studied associations of several reported ENT symptoms and chronic rhinosinusitis (CRS)—defined using patient-reported information and examination findings—with reported sputum production and shortness of breath—using ordinal logistic regression. In a subgroup with available lung function results, we used linear regression to study associations of CRS and FEV_1_, accounting for relevant factors.

**Results:** We included 457 patients [median age: 15; interquartile range (IQR) 10–24; 54% males]. Shortness of breath associated with reported nasal symptoms and ear pain of any frequency, often or daily hearing problems, headache when bending down [odds ratio (OR) 2.1; 95% confidence interval (CI) 1.29–3.54], and CRS (OR 2.3; 95% CI 1.57–3.38) regardless of polyp presence. Sputum production associated with daily reported nasal (OR 2.2; 95% CI 1.20–4.09) and hearing (OR 2.0; 95% CI 1.10–3.64) problems and CRS (OR 2.1; 95% CI 1.48–3.07). We did not find any association between CRS and FEV_1_.

**Conclusion:** Reported upper airway symptoms and signs of CRS associated with reported pulmonary symptoms; however, not with lung function. Our results emphasise assessing and managing upper and lower respiratory disease as a common, interdependent entity among patients with PCD.

## Introduction

Nearly all patients with primary ciliary dyskinesia (PCD) report chronic nasal problems caused by poor mucociliary clearance, leading to mucus stagnation in upper and lower airways [1–3]. Clogged airways facilitate recurrent infections, chronic microbial colonisation, and airway inflammation, leading further to chronic rhinosinusitis (CRS) and bronchiectasis [4, 5]. In other respiratory diseases, such as asthma and cystic fibrosis (CF), evidence supports the theory of “the unified airway” [6, 7]. Published studies highlighted the association of CRS with chronic obstructive pulmonary disease (COPD) [8, 9]. However, for PCD upper and lower airway manifestations are often managed independently. A common approach is usually considered when treatments for pulmonary exacerbations fail and sinuses become considered possible reservoirs for pulmonary colonisation, chronic lung infections, and lung function deterioration [10].

So far, only a few single-centre studies attempted connecting the dots between upper and lower airways in PCD [11–13]. A study in a small cohort in Denmark presented simultaneous infections of the sinuses and lower airways with the same pathogen among patients with PCD [10]. A French study assessed associations between ear-nose-throat (ENT) symptoms and lung function among adult patients with PCD and reported otitis media with effusion associated with airway obstruction [forced expiratory volume (FEV_1_) < 70%] [14]. Otherwise, scarce evidence exists about possible associations of sinonasal and otologic symptoms and signs of disease with pulmonary symptoms in PCD and whether patients with more upper airways symptoms also have more advanced lung disease. Since it remains unclear what—if any—upper respiratory characteristics possibly associate with reduced lung function, we studied associations 1) between patient-reported upper and lower respiratory symptoms; 2) between CRS—with or without nasal polyps—and reported lower respiratory symptoms; and 3) between CRS—with and without nasal polyps—and lung function.

## Methods

### Study design and population

We analysed cross-sectional data from our ENT prospective, international cohort of patients with PCD (EPIC-PCD)—the first PCD cohort focused on upper respiratory disease [15]. EPIC-PCD started recruiting patients with PCD in February 2020, following them during regular ENT visits at participating centres. We nested examinations in regular care and collected additional questionnaires. For this study, we included eligible patients with data entered in the EPIC-PCD database by May 15, 2023 from 13 participating centres (Amsterdam, Ankara, Berlin, Bern, Cyprus, Istanbul, Leuven, Liège, Münster, Oslo, Paris, Southampton, Valencia) in 10 countries. We included participants of all ages with PCD with ENT examination and completed symptom questionnaire within a two week interval of the examination.

The EPIC-PCD study is hosted at the University of Bern (clinicaltrials.gov identifier: NCT04611516). We received ethical approval from each participating centre and ethics committee for human research in accordance with local legislation. We obtained informed consent or assent from either participants or parents or caregivers of participants 14 years or younger as previously described [3, 16]. Our reporting conforms with the Strengthening the Reporting of Observational studies in Epidemiology (STROBE) statement [17].

### Patient-reported symptoms

We collected patient-reported symptoms using the disease-specific FOLLOW-PCD questionnaire (version 1.0), which is part of the standardised PCD-specific form FOLLOW-PCD developed for collecting clinical information for research and clinical follow-up [18]. The FOLLOW-PCD questionnaire was designed with three versions for three age groups: adults, adolescents 14–17 years, and parents or caregivers of children with PCD younger than 14 years. It is available in the local languages of all participating centres. Symptom-related questions asked about frequency and characteristics of symptoms during the previous three months. For the upper respiratory symptoms, we focused on chronic nasal symptoms, headache when bending down as proxy for sinusitis, ear pain, and hearing problems. For lower respiratory symptoms we focused on shortness of breath and sputum production, which included any reported cough with expectorated or swallowed secretions. Symptom frequency options included daily, often, sometimes, rarely, and never (five-point Likert scale). The questionnaire also included questions about health-related behaviours, such as smoking exposure and living conditions, during the past 12 months. Depending on available response categories, we recoded missing answers as “unknown,” “no,” or “never.”

### Clinical examinations

ENT specialists performed routine examinations—sinonasal examinations by nasal endoscopy or anterior rhinoscopy if tolerated by participants, otoscopy, tympanometry, and audiometry among others—at planned consultations, according to local protocols. Examination findings were recorded in a standardised way using the ENT examination module of the FOLLOW-PCD form [18]. If spirometry was performed before or after 1 month from ENT consultation, we also recorded FEV_1_ (Forced Expiratory Volume in 1 Second) values. Participating centres performed spirometry according to American Thoracic Society/European Respiratory Society (ERS) guidelines [19] during routine planned visits and not at exacerbation or during respiratory tract infection. We calculated FEV_1_ z-scores based on the Global Lung Initiative 2021 reference values [20]. We calculated body mass index (BMI) using height and weight reported at ENT or spirometry visit date. For adults, we classified BMI as underweight (<18.5 kg/m ^2^), normal (≥18.5 to <25), pre-obesity (≥25 to <30), obesity class I (≥30 to <35), obesity class II (≥35 to <40), or obesity class III (≥40) by World Health Organization (WHO) standards [21]. For children and adolescents younger than 18 years, we calculated sex and age-specific BMI z-scores and categorised by thinness (<-2 z-scores), normal (−2 to 1 z-scores), overweight (1 to 2 z-scores), and obesity (>2 z-scores) based on 2007 WHO references [22].

### Definition of CRS

We created a composite exposure variable CRS to study CRS associations—with and without polyps—with reported lower respiratory symptoms and lung function. The dichotomous composite variable included 1) daily or often reported nasal symptoms and 2) examination findings of nasal discharge (sero-mucous, muco-purulent, or mixed with blood) or nasal oedema at examination.

### Diagnosis and other clinical information from charts

Participants were diagnosed at participating centres following ERS guidelines [23] as described in previous publications [3, 16]. Ultrastructural defects were categorised based on the international consensus guideline for reporting transmission electron microscopy (TEM) results, which defined class 1 (hallmark, name outer dynein arm defects, outer and inner dynein arm defects, and microtubular disorganisation with inner dynein arm defects) and class 2 defects, such as central complex defects [24]. Further data collected included information on laterality defects and prescribed medication for upper and lower airways. We entered all collected data in the Research Electronic Data Capture study database based on the FOLLOW-PCD modules [18].

### Statistical analysis

We described population characteristics and patient or parent-reported symptoms for the total population and separately among age groups 0–6, 7–14, 15–30, 31–50, and older than 50 years. For continuous variables, we used median and interquartile range (IQR). For categorical variables, we used numbers and proportions, and we compared differences between age groups using Pearson’s Chi square and Kruskal-Wallis rank test. We studied possible associations of reported frequency of nasal symptoms, ear pain, hearing problems, and of reported headache when bending down with reported frequency of shortness of breath and sputum production using multivariable ordinal logistic regression, adjusting for age and sex. In separate multivariable ordinal logistic regression models, we assessed association of CRS (as defined above) with frequency of shortness of breath and sputum production, adjusting for factors possibly associated with respiratory disease such as age, sex, age at diagnosis, nasal polyp status, and smoking status as either active, passive or no tobacco smoke exposure.

For a subgroup of patients with available FEV_1_, we assessed the association of CRS with FEV_1_ z-score using linear regression and adjusting for age, sex, nasal polyps, smoking status, and prescribed nasal corticosteroids, prophylactic antibiotics, nasal rinsing, and inhaled corticosteroids. For all models, we chose factors based on data availability and clinical importance to the study team. We noted a collinearity of age and age at diagnosis, so it was not possible to include both in our main model. Since separate models showed similar results, we only included age. Among a subgroup of participants with available TEM results, we repeated our regression models, including age and category of ciliary ultrastructural defect to study whether ciliary ultrastructural defect was a risk factor for a possible association between CRS and lower airway symptoms or FEV_1_. We performed all analyses with Stata version 15 (StataCorp LLC, Texas, USA).

## Results

### Study population

By mid-May 2023, 504 (85%) of 596 invited patients enrolled into the EPIC-PCD study (Figure 1). Of them, 457 had data entered in the database and fulfilled eligibility criteria. We included 286 (63%) children and 171 (37%) adults; 54% male (table 1); median age 15 (interquartile range: IQR 10–24); 162 (35%) with situs inversus totalis; and 36 (8%) cardiovascular malformation—five with severe malformation, such as transposition of the great arteries. Height and weight were available for about 80% of the study population, half with normal BMI (table 1). Obesity class III was more prevalent among adults 31 years and above. We did not find any differences by sex for any of these characteristics. We present a summary of sinonasal examination findings used to define CRS and prescribed treatments in Table S2.

**Figure 1:**
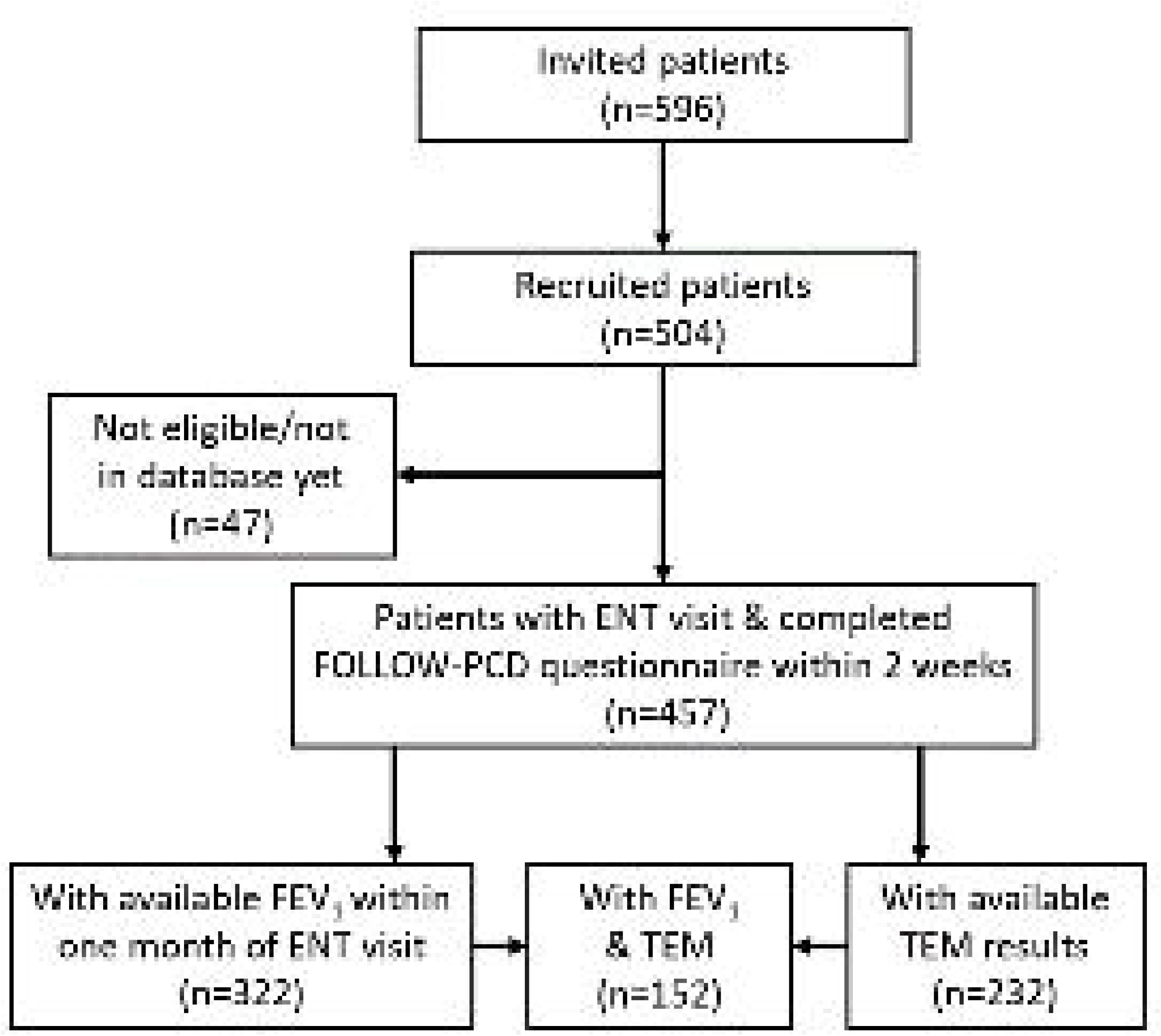
Flowchart of people who participated in EPIC-PCD

**Table 1:** Characteristics of EPIC-PCD participants, overall and by age group (N=457)

|  | **Total**<br>**N (%)** | **Age 0–6 y**<br>**N (%)** | **Age 7–14 y**<br>**N (%)** | **Age 15–30 y**<br>**N (%)** | **Age 31–50 y**<br>**N (%)** | **Age >50 y**<br>**N (%)** | **p-value<sup>a</sup>** |
| --- | --- | --- | --- | --- | --- | --- | --- |
| **Number of participants** | 457 (100) | 47 (100) | 149 (100) | 173 (100) | 54 (100) | 34 (100) |  |
| **Age**<br>median (IQR) | 15 (10–24) | 2 (4–5) | 10 (8–12) | 18 (16–22) | 38 (34–42) | 57 (55–63) |  |
| **Male sex** | 246 (54) | 26 (55) | 82 (55) | 90 (52) | 30 (56) | 18 (53) | 0.980 |
| **Age of PCD diagnosis**<br>median (IQR) | 9 (4–18) | 0 (0–2) | 6 (1–8) | 13 (9–17) | 34 (29–37) | 51 (44–56) |  |
| **Consanguinity** |  |  |  |  |  |  | <0.001 |
| Yes | 124 (27) | 7 (15) | 46 (31) | 56 (32) | 11 (20) | 4 (12) |  |
| No | 167 (37) | 15 (32) | 50 (33) | 67 (39) | 27 (50) | 8 (23) |  |
| Not reported | 166 (36) | 25 (53) | 53 (36) | 50 (29) | 16 (30) | 22 (65) |  |
| **Situs** |  |  |  |  |  |  | 0.001 |
| Situs inversus totalis | 162 (35) | 26 (55) | 56 (37) | 64 (37) | 9 (17) | 7 (21) |  |
| Situs ambiguous | 5 (1) | 0 (0) | 2 (1) | 3 (2) | 0 (0) | 0 (0) |  |
| Situs solitus | 278 (61) | 20 (43) | 88 (60) | 105 (60) | 40 (74) | 25 (73) |  |
| Not reported | 12 (3) | 1 (2) | 3 (2) | 1 (1) | 5 (9) | 2 (6) |  |
| **Cardiovascular malformation** |  |  |  |  |  |  | 0.003 |
| Yes | 36 (8) | 7 (15) | 13 (9) | 14 (8) | 2 (4) | 0 (0) |  |
| No | 328 (72) | 31 (66) | 116 (78) | 125 (72) | 37 (68) | 19 (56) |  |
| Not reported | 93 (20) | 9 (19) | 20 (13) | 34 (20) | 15 (28) | 15 (44) |  |
| **Active smoking** |  |  |  |  |  |  | <0.001 |
| Yes, daily | 4 (1) | NA | NA | 3 (2) | 0 (0) | 1 (3) |  |
| Yes, rarely | 6 (1) | NA | NA | 3 (2) | 2 (4) | 1 (3) |  |
| Ex-smoker | 17 (4) | NA | NA | 2 (1) | 10 (18) | 5 (15) |  |
| Never smoker | 222 (49) | NA | NA | 156 (90) | 40 (74) | 26 (76) |  |
| Not reported | 208 (45) | NA | NA | 9 (5) | 2 (4) | 1 (3) |  |
| **Smoking in household** |  |  |  |  |  |  | <0.001 |
| Yes | 83 (18) | 7 (15) | 29 (20) | 38 (22) | 5 (9) | 4 (12) |  |
| No | 298 (65) | 36 (77) | 109 (73) | 98 (56) | 36 (67) | 19 (56) |  |
| Not reported | 76 (17) | 4 (8) | 11 (7) | 37 (21) | 13 (24) | 11 (32) |  |

EPIC-PCD: Ear-nose throat prospective international cohort of patients with primary ciliary dyskinesia. y: years. Characteristics are presented as N and column %, age as median and IQR: interquartile range. NA: not applicable; age-specific questionnaire version does not include question on active smoking in this age category. <sup>a</sup>Chi-square test of independence.

**Table 1 (continued):** Characteristics of EPIC-PCD participants, overall and by age group (N=457)
|  | **Total**<br>**N (%)** | **Age 0–6 y**<br>**N (%)** | **Age 7–14 y**<br>**N (%)** | **Age 15–30 y**<br>**N (%)** | **Age 31–50 y**<br>**N (%)** | **Age >50 y**<br>**N (%)** | **p-value<sup>a</sup>** |
| --- | --- | --- | --- | --- | --- | --- | --- |
| **Number of participants** | 457 (100) | 47 (100) | 149 (100) | 173 (100) | 54 (100) | 34 (100) |  |
| **BMI [kg/m<sup>2</sup>]**<br>**mean (IQR)<sup>b</sup>** |  |  |  | 21.1<br>(19.7–24.1) | 23.2<br>(20.8–27.4) | 28.1<br>(21.6–35.3) | <0.001<sup>d</sup> |
| **BMI z-score**<br>**median (IQR)<sup>c</sup>** |  | -0.5<br>(-1.3–0.4) | -0.06<br>(-0.9–1.2) | 0.1<br>(-0.9–1.0) |  |  | 0.402<sup>d</sup> |
| **BMI categories** |  |  |  |  |  |  | <0.001 |
| Thinness/<br>underweight | 26 (6) | 1 (2) | 7 (4) | 12 (7) | 5 (9) | 1 (3) |  |
| Normal<br>weight | 225 (49) | 6 (13) | 83 (56) | 107 (62) | 23 (43) | 6 (17) |  |
| Pre-obesity/<br>overweight | 66 (14) | 1 (2) | 22 (15) | 25 (14) | 13 (24) | 5 (15) |  |
| Obese/obesity<br>class I | 24 (5) | 0 (0) | 9 (6) | 8 (5) | 3 (6) | 4 (12) |  |
| Obesity<br>class II | 7 (2) | 0 (0) | 0 (0) | 3 (2) | 0 (0) | 4 (12) |  |
| Obesity<br>class III | 31 (7) | 0 (0) | 0 (0) | 7 (4) | 10 (18) | 14 (41) |  |
| Missing | 78 (17) | 39 (83) | 28 (19) | 11 (6) | 0 (0) | 0 (0) |  |
| **FEV<sub>1</sub> z-score**<br>**median (IQR)** | -1.9<br>(-2.9 to -0.6) | -0.7<br>(-2.0 to -0.3) | -1.6<br>(-2.5 to 0.4) | -2.1<br>(-3.0 to 0.8) | -2.3<br>(-3.3 to -0.8) | -1.7<br>(-3.6 to -0.6) | 0.008<sup>d</sup> |

EPIC-PCD: Ear-nose throat prospective international cohort of patients with primary ciliary dyskinesia. y: years. Characteristics are presented as N and column %. <sup>a</sup>Chi-square test of independence. <sup>b</sup>BMI (Body mass index) of 145 adults (≥18 years) based on World Health Organization (WHO) standards. <sup>c</sup>BMI z-score from 208 children (<18 years) based on WHO standards. FEV<sub>1</sub> z-score available from 322 participants. <sup>d</sup>Kruskal–Wallis test.

### Association of reported upper and lower respiratory symptoms

Reported upper and lower respiratory symptoms were common for all age groups, especially nasal symptoms and sputum production (Table S1). We found reported frequency of shortness of breath increased with any reported frequency of nasal symptoms [odds ratio (OR): 4.2; 95% confidence interval (CI): 2.18–8.16 for daily nasal symptoms compared with no nasal symptoms] and with reported headache when bending down (OR: 2.1; 95% CI 1.29–3.54) (Figure 2). For frequency of sputum production, we only found evidence of association with daily nasal symptoms (OR: 2.1; 95% CI 1.20–4.09). Regarding reported ear symptoms, any frequency of ear pain (OR: 3.7; 95% CI 1.62– 8.44 for daily ear pain compared with no ear pain) and daily (OR: 2.0; 95% CI 1.13–3.71) or often (OR: 1.9, 95% CI 1.10–3.46) reported hearing problems compared with no hearing problems associated with frequency of shortness of breath (Figure 3). Reported daily hearing problems also associated with higher frequency of sputum production (OR: 2.0; 95% CI 1.10–3.64). Male sex was less likely associated with shortness of breath (Figures 2–3; OR for male sex in all models: 0.7; 95% CI ranged from 0.50–0.97 to 0.52–1.07). We found no differences by age.

**Figure 2:**
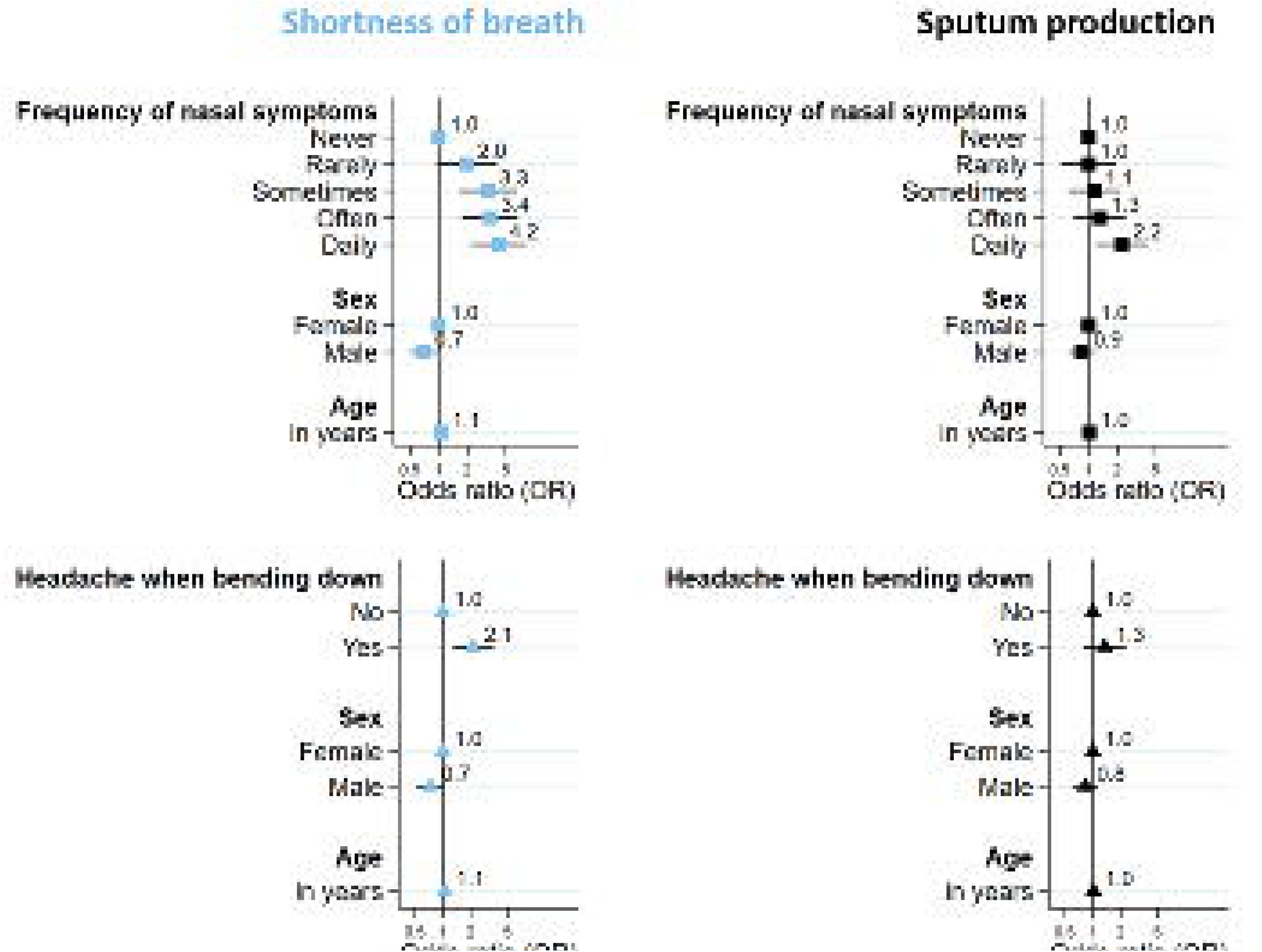
Association of patient-/parent-reported nasal symptoms or headache when bending down with shortness of breath and sputum production among EPIC-PCD participants (N=457).

**Figure 3:**
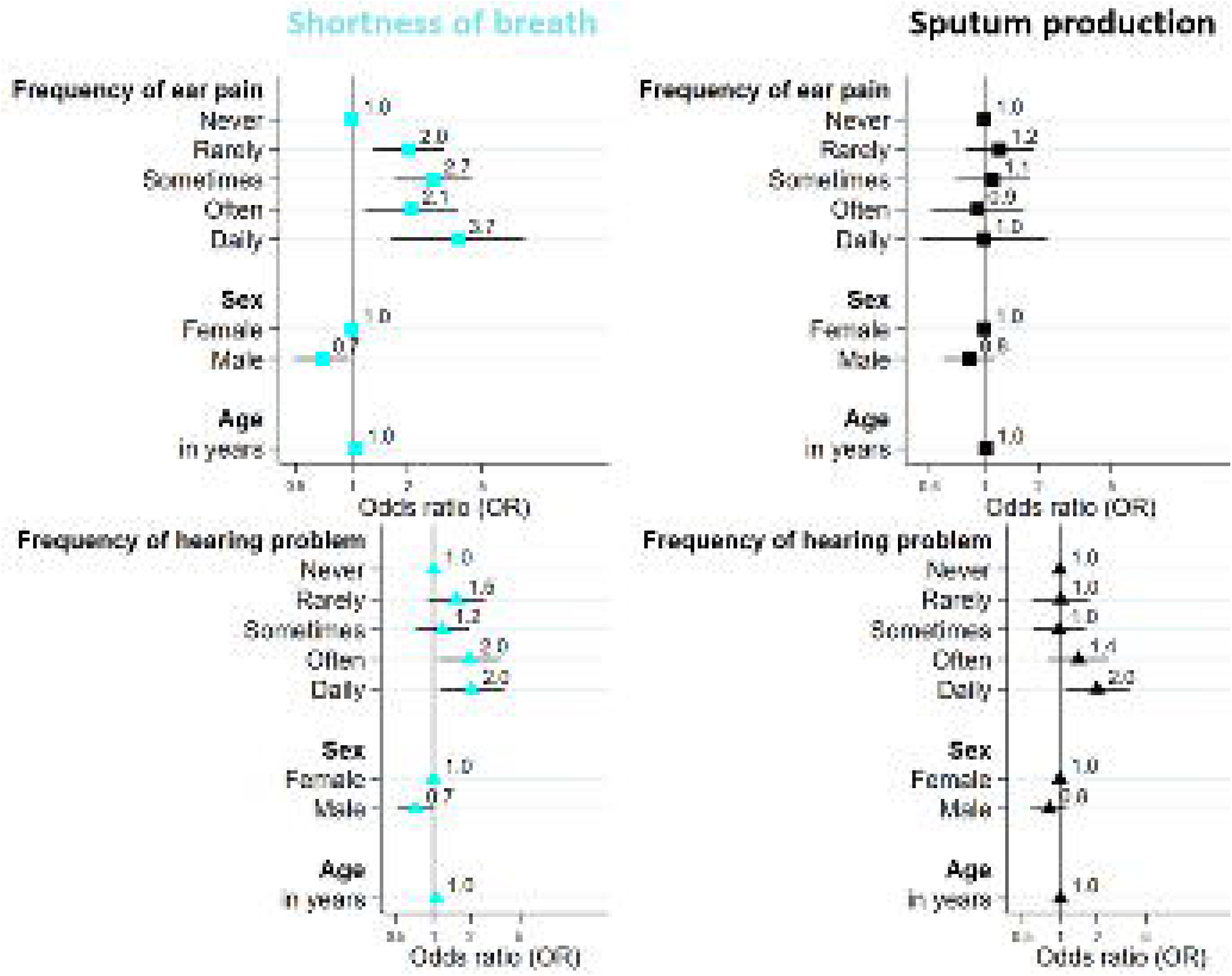
Association of patient-/parent-reported ear pain and hearing problems with shortness of breath and sputum production among EPIC-PCD participants (N=457).

### Association of CRS with lower respiratory symptoms

We found evidence of association between CRS and reported frequency of shortness of breath (OR 2.3; 95% CI 1.53–3.32) and sputum production (OR 2.1; 95% CI 1.48–3.06) (Figure 4). We did not find any differences related to the presence or absence of nasal polyps accompanying CRS, tobacco smoke exposure, or by age or sex for both lower respiratory symptoms. Among 232 participants with available TEM results (Table S3), we found no difference in reported shortness of breath and sputum production by ciliary ultrastructural defect class (Table S4).

**Figure 4:**
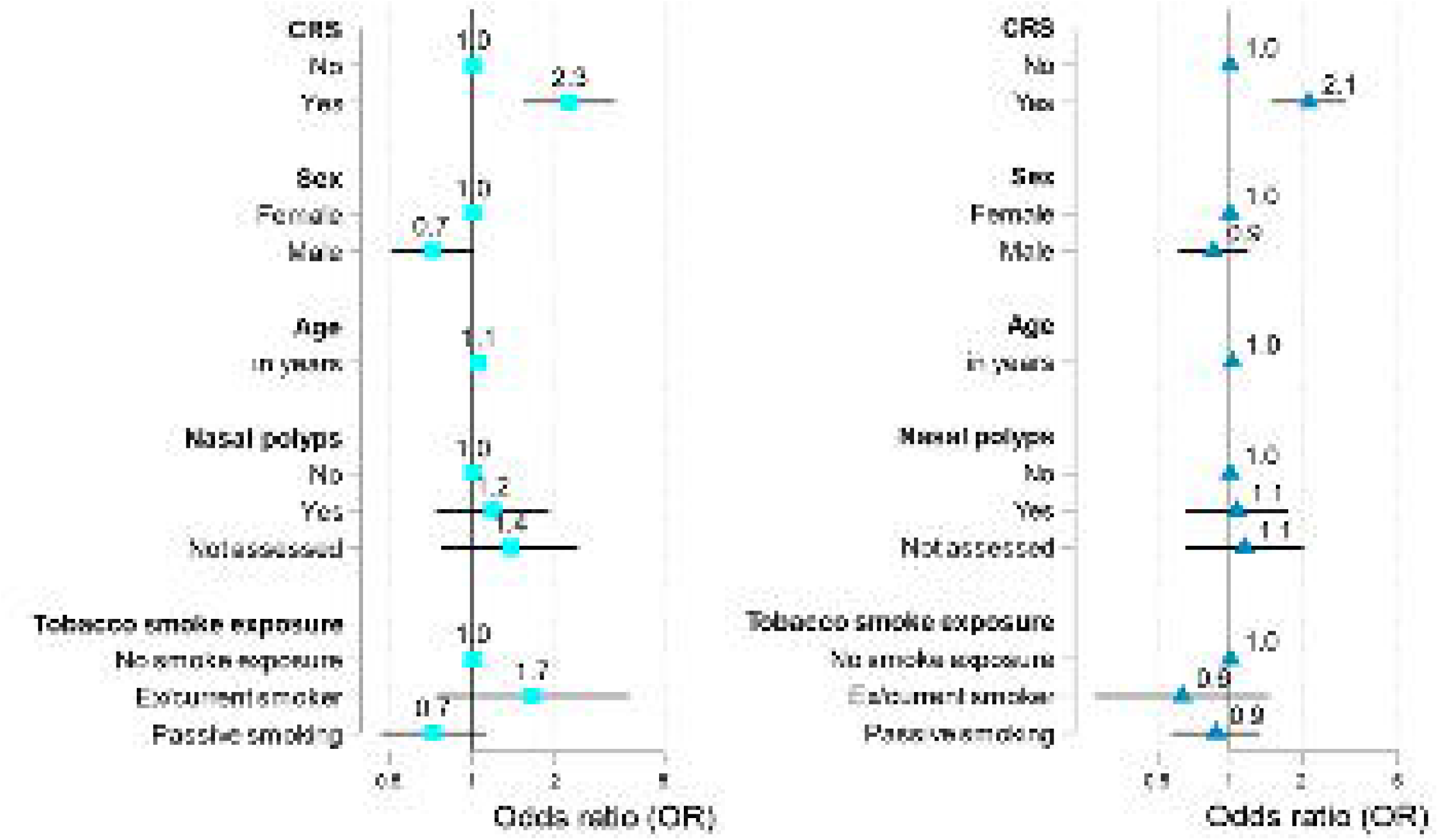
Association of chronic rhinosinusitis (CRS) with patient-/parent-reported shortness of breath and sputum production among EPIC-PCD participants (N=457).

### Association of CRS with lung function

Within one month of ENT visit, 322 participants had available spirometry with a median F_1_EV z-score of −1.90 (IQR −2.9 to −0.6) (Table 1). We found no association between CRS, independently of polyps and FEV_1_ z-score, adjusting for possible confounders (Table 2). Participants prescribed nasal corticosteroids showed higher FEV_1_ z-scores (OR 2.26; 95% CI 0.44–4.07). We also found higher FEV_1_ z-score among participants not prescribed inhaled corticosteroids (OR 5.54; 95% CI 2.73–8.34). In a subgroup of 152 participants with available TEM results and spirometry, we found no evidence of association of CRS with FEV_1_ z-score and no differences by defect (Table S5).

**Table 2:** Association of chronic rhinosinusitis (CRS) with FEV_1_ z-score among EPIC-PCD participants (N=322)

|  | Coefficient | 95% confidence interval | p-value |
| --- | --- | --- | --- |
| **CRS** | 0.80 | -0.83 – 2.24 | 0.367 |
| **Age** | -0.02 | -0.09–0.04 | 0.470 |
| **Male sex** | 0.01 | -1.37–1.39 | 0.988 |
| **Nasal polyps** |  |  |  |
| Yes | 0.58 | -1.43–2.60 | 0.568 |
| Not assessed | -1.13 | -3.80–1.55 | 0.408 |
| **Smoking** |  |  |  |
| Ex/current smoker | 7.44 | 3.27–11.62 | 0.001 |
| Passive smoking | 0.08 | -1.70–1.85 | 0.933 |
| **Nasal corticosteroids** |  |  |  |
| Yes | 2.26 | 0.44–4.07 | 0.015 |
| Not described | -1.85 | -5.00–1.31 | 0.250 |
| **Nasal rinsing** |  |  |  |
| Yes | -1.50 | -3.15–0.16 | 0.077 |
| Not described | -1.92 | -4.26–0.42 | 0.107 |
| **Inhaled corticosteroids** |  |  |  |
| Yes | -0.34 | -1.93–1.26 | 0.667 |
| Not described | 5.54 | 2.73–8.34 | <0.001 |
| **Prophylactic antibiotic** |  |  |  |
| Yes | -0.60 | -2.24–1.04 | 0.473 |
| Not described | -2.93 | -5.34 – -0.53 | 0.017 |

Results of linear regression model.
EPIC-PCD: Ear-nose throat prospective international cohort of patients with primary ciliary dyskinesia. CRS – chronic rhinosinusitis composite exposure variable consisting of daily or often reported nasal symptoms and examination findings of nasal discharge (sero-mucous, muco-purulent, or mixed with blood) or nasal oedema.

## Discussion

Our results showed an association between upper respiratory disease and reported lower respiratory symptoms. Particularly, shortness of breath associated with reported nasal symptoms and ear pain of any frequency; often or daily hearing problems; headache when bending down and with CRS—defined using patient-reported information and examination findings—regardless of polyp presence. Sputum production associated with daily reported nasal symptoms and hearing problems, as well as CRS again regardless of polyp presence. Contrary to symptom findings, we did not find any association between CRS and reduced lung function measured by spirometry.

### Strengths and limitations

EPIC-PCD is the first prospective, international ENT cohort for PCD and—to our knowledge— the first study combining patient-reported symptoms and findings from clinical examinations of upper and lower airways. We followed participants during regular visits using FOLLOW-PCD modules, which makes collecting standardised data possible for all participating centres. Since invited patients were interested and it required little effort on their part, most agreed to participate in the study. Not all participants met inclusion criteria (ENT examination and completed questionnaire). We believe exclusion was at random and mostly based on personnel resources or organisational issues at participating centres. However, it is possible participants with fewer symptoms were less likely to complete questionnaires and fulfil inclusion criteria, introducing selection bias. Lung function measurements were unavailable for some participants within one month of study visit, yet entirely dependent on participating centres—several countries organise pulmonary and ENT visits separately. Since questions about symptoms involved the previous three months, we expect minimal risk of recall bias. EPIC-PCD started recruitment during the SARS-CoV-2 pandemic; however, infection incidence was low among people with PCD [25]. We expect possible lower reporting of symptoms, especially in the beginning of the study likely from shielding behaviour [26]. Despite this being the largest study of its kind, we still lack statistical power to consider several other possible factors possibly influencing associations between upper and lower airways disease, including comorbidities, such as asthma and information about management.

### Comparison with other studies

Few studies assessed associations between upper and lower respiratory disease in PCD and none used standardised information on symptoms to assess possible associations. A French study evaluating sinonasal disease among 64 adults with PCD found otitis media with effusion associated with FEV_1_ <70% [14]. We tested for possible association of CRS with FEV_1_ and found no evidence of association. However, we found patient-reported ear pain and hearing problems associated with lower respiratory symptoms, specifically shortness of breath and sputum production. A smaller single-centre study showed simultaneous infections with the same pathogens in sinuses and lungs of patients in Denmark who underwent sinus surgery [10]. The concept of “the unified airway” in PCD was further supported by another Danish study showing the same *Pseudomonas aeruginosa*clone in sinuses and lungs [27]. Unfortunately, our observational study rarely included simultaneous upper and lower airway cultures since they are only routinely collected at few participating centres, so we could not use microbiology information to support our findings further.

Studies on other chronic lung diseases showed similar associations between upper and lower respiratory disease [28, 29]. A large population-based study among adults in southern Sweden found nasal symptoms frequently coexisted with both self-reported diagnoses of asthma and chronic bronchitis/emphysema—only 33% of the total population reported nasal symptoms compared with 40% among participants with self-reported COPD—suggesting pan-airway engagement as common for both diseases [30]. A Canadian study with 121 participants diagnosed with CF compared FEV_1_ (% predicted value) between individuals with and without CRS and found no difference (mean difference, 2.0%; 95% CI, −8.1% to 13.0%), which is similar to our study [31].

### Interpretation of findings

Our findings support the concept of “the unified airway” in PCD, particularly the association between nasal symptoms and CRS and lower respiratory symptoms—a finding possibly explained by increased mucus production or decreased mucosal clearance along “the unified airway.” Since ciliary function is affected in upper and lower airways, we expect patients with PCD to report symptoms from both and account for any differences based on disease severity. Interactions with allergic rhinitis might be possible; however, lack of detailed data on most participants precluded examining such a hypothesis. We believe the interactions as small since we previously found no associations between sinonasal disease and any particular season, especially not pollination seasons [3]. For some patients, coexisting posterior nasal drip explained the association of daily nasal symptoms with sputum production. Recently, heterogeneity of clinical phenotypes in PCD stimulated much discussion [32–34]. Any evidence of association we found between ear symptoms and shortness of breath could also be explained by possible underlying CRS in these patients. Ear pain and hearing problems might be symptoms of Eustachian tube dysfunction, which is prevalent among patients with CRS [35, 36]. Our study suggests upper and lower respiratory symptoms occur dependently for most patients with PCD. Therefore, it is probable differences in upper and lower airway disease between PCD clinical phenotypes mainly relate to disease severity and less to prevalence of specific respiratory symptoms.

We found associations of CRS with lower respiratory symptoms yet not with FEV_1_ measured by spirometry. Although spirometry is the most commonly used method for pulmonary assessment for PCD [37], it appears not sensitive enough for patients with PCD, particularly children [38]. It is prone to large intra-individual variability, which complicates assessing possible associations. Lung disease in PCD is complex and cannot be assessed only with spirometry as there is often discordance between lung function and impairment shown on imaging modalities [39]. Other tests such as multiple breath washout appear more sensitive than spirometry for detecting pulmonary disease [40–42]; we recommend studying associations using these measurements.

## Conclusion

Our study shows reported upper airway symptoms and examination findings of CRS associated with reported lower respiratory symptoms; however, not with airway obstruction assessed by lung function. Upper and lower airway disease occurs interdependently; to improve clinical outcomes for patients with PCD, it needs assessing and managing as a common entity with appropriate clinical and patient-reported measures.

## Availability of data and materials

Upon reasonable request, our datasets for the present study are available from the study PI, Dr. Myrofora Goutaki. The EPIC - PCD dataset includes individual patient data of people with a rare disease. Although data are pseudonymised, data possibly still include sensitive information which possibly lead to identifying participants; therefore, participants were not asked to consent having their data deposited or shared publicly.

## Author contribution

M Goutaki developed the concept and design of the study. M Goutaki and YT Lam managed the study. YT Lam cleaned, standardised the data, and performed statistical analyses supervised by M Goutaki. YT Lam and M Goutaki drafted the manuscript. All authors commented and revised the manuscript. YT Lam and M Goutaki take final responsibility for content.

## Statement on funding sources and conflicts of interest

The Swiss National Science Foundation Ambizione fellowship (PZ00P3_185923) funded the study. Authors participate in the BEAT-PCD (Better experimental approaches to treat PCD) clinical research collaboration—supported by the European Respiratory Society—and most centres are members of the PCD core of ERN-LUNG (European Reference Network on rare respiratory diseases). JF Papon reports personal fees from Sanofi, GSK, Medtronic, and ALK outside submitted work. M Alexandru received personal fees from Sanofi and ALK outside of submitted work. M Boon reports grants from Forton grant (King Baudouin Foundation) 2020-J1810150-217926 cystic fibrosis research and personal fees from Vertex outside submitted work. N Lorent received honoraria to her institution from GSK, INSMED, AN2 Therapeutics outside submitted work and a travel grant from Pfizer. J Roehmel received grants, clinical study recompensations from Vertex, INSMED, and Medical Research Council/UK, BMBF, Mukoviszidose Institut outside of submitted work.

## Acknowledgements

We thank all people with PCD and their families participating in EPIC-PCD and PCD support organisations (especially PCD Family Support Group UK; Association ADCP France; Kartagener Syndrom und Primäre Ciliäre Dyskinesie e. V. Deutschland/ Deutschschweiz; Asociación Nacional de Pacientes con Discinesia Ciliar Primaria DCP España/PCD Spain) for their close collaboration. We also thank all researchers of the participating centres involved in enrolment, data collection, and data entry who work closely with us (listed below as collaborators of the EPIC-PCD study). We are grateful for everyone who contributed to translations of the FOLLOW-PCD questionnaire in Dutch, Flemish, French, Norwegian, Spanish, and Turkish. Lastly, we thank Kristin Marie Bivens (ISPM, University of Bern) for her editorial assistance.

*EPIC-PCD team (listed in alphabetical order): Dilber Ademhan (Hacettepe University, Turkey), Mihaela Alexandru (AP-HP, France), Andreas Anagiotos (Nicosia General Hospital, Cyprus), Miguel Armengot (La Fe University and Polytechnic Hospital, Spain), Lionel Benchimol (University Hospital of Liège, Belgium), Achim G Beule (University of Münster, Germany), Irma Bon (Vrije Universiteit, the Netherlands), Mieke Boon (University Hospital Leuven, Belgium), Marina Bullo (University of Bern, Switzerland), Andrea Burgess (University of Southampton, UK), Doriane Calmes (University Hospital of Liège, Belgium), Carmen Casaulta (University of Bern, Switzerland), Marco Caversaccio (University of Bern, Switzerland), Nathalie Caversaccio (University of Bern, Switzerland), Bruno Crestani (RESPIRARE, France), Suzanne Crowley (University of Oslo, Norway), Sinan Ahmed. D. Dheyauldeen (University of Oslo, Norway), Sandra Diepenhorst (Vrije Universiteit, The Netherlands), Nagehan Emiralioglu (Hacettepe University, Turkey), Ela Erdem Eralp (Marmara University, Turkey), Pinar Ergenekon (Marmara University, Turkey), Nathalie Feyaerts (University Hospital Leuven, Belgium), Gavriel Georgiou (Nicosia General Hospital, Cyprus), Amy Glen (University of Southampton, UK), Christine van Gogh (Vrije Universiteit Amsterdam, the Netherlands), Yasemin Gokdemir (Marmara University, Turkey), Myrofora Goutaki (University of Bern, Switzerland), Onder Gunaydın (Hacettepe University, Turkey), Eric G Haarman (Vrije Universiteit Amsterdam, The Netherlands), Amanda Harris (University of Southampton, UK), Lilia Marianne Hartung (Charité-Universitätsmedizin Berlin, Germany), Isolde Hayn (Charite-Universitatsmedizin Berlin, Germany), Simone Helms (University of Münster, Germany), Sara-Lynn Hool (University of Bern, Switzerland), Isabelle Honoré (RESPIRARE, France), Isabel Ibáñez (La Fe University and Polytechnic Hospital, Spain), Hasnaa Ismail Koch (University of Southampton, UK), Bülent Karadag (Marmara University, Turkey), Céline Kempeneers (University Hospital of Liège, Belgium), Synne Kennelly (University of Oslo, Norway), Elisabeth Kieninger (University of Bern, Switzerland), Sookyung Kim (AP-HP, France), Panayiotis Kouis (University of Cyprus, Cyprus), Yin Ting Lam (University of Bern, Switzerland), Philipp Latzin (University of Bern, Switzerland), Marie Legendre (RESPIRARE, France), Natalie Lorent (University Hospital Leuven, Belgium), Jane S Lucas (University of Southampton, UK), Bernard Maitre (RESPIRARE, France), Alison McEvoy (University of Southampton, UK), Rana Mitri-Frangieh (RESPIRARE, France), David Montani (RESPIRARE, France), Loretta Müller (University of Bern, Switzerland), Noelia Muñoz (La Fe University and Polytechnic Hospital, Spain), Heymut Omran (University of Münster, Germany), Ugur Ozcelik (Hacettepe University, Turkey), Beste Ozsezen (Hacettepe University, Turkey), Samantha Packham (University of Southampton, UK), Jean-François Papon (AP-HP, France), Clara Pauly (University Hospital of Liège, Belgium), Charlotte Pioch (Charité-Universitätsmedizin Berlin, Germany), Anne-Lise ML Poirrier (University Hospital of Liège, Belgium), Johanna Raidt (University of Münster, Germany), Ana Reula (La Fe University, Spain), Rico Rinkel (Vrije Universiteit Amsterdam, the Netherlands), Jobst Roehmel (Charité-Universitätsmedizin Berlin, Germany), Andre Schramm (University of Münster, Germany), Catherine Sondag (University Hospital of Liège, Belgium), Simone Tanner (Vrije Universiteit, the Netherlands), Nicoletta Tanou (University of Cyprus, Cyprus), Guillaume Thouvenin (RESPIRARE, France), Woolf T Walker (University of Southampton, UK), Hannah Wilkins (University of Southampton, UK), Panayiotis Yiallouros (University of Cyprus, Cyprus), Ali Cemal Yumusakhuylu (Marmara University, Turkey), Niklas Ziegahn (Charité-Universitätsmedizin Berlin, Germany)

## Notes

### Author Declarations

The study has been reviewed and approved by the local Human Research Ethics Committees at every participating centre, based on local legislation. We list below the names of the ethics committees which approved the study and the approval reference numbers, when applicable. A. The following centres have a pre-existing or new ethical approval, which allows the contribution of pseudonymised data to observational collaborative international studies (covers the EPIC-PCD study): 1. University Childrens Hospital Charite-Universitaetsmedizin, Berlin, Germany: Ethical Committee Charite (EA2/003/21) 2. University Childrens hospital, Bern, Switzerland: Cantonal Ethics Committee of Bern (KEKBE: 060/2015) 3. University of Cyprus: Ethical Committee for biomedical research in Leukosia Cyprus (EEBK/EΠ/2013/21) 4. Marmara University Istanbul, Turkey: Ethical Committee of Marmara University (09.2018.395) 5. University Hospital of Southampton, United Kingdom: Southampton and South West Hampshire research ethics committee (06/Q1702/109) 6. University hospital Muenster, Germany: Ethical committee (2011-270-f-S) B. The following centres applied for ethical approval to participate specifically to the EPIC-PCD study: 1. VU University medical center (VUmc), Amsterdam, The Netherlands: The Medical Ethics Review Committee of VU University Medical Center reviewed the application and concluded on 24th of November 2020 that no approval is needed to participate to the EPIC-PCD cohort as the Medical Research Involving Human Subjects Act does not apply to the study. 2. Hacettepe University, Ankara, Turkey: Non-interventional clinical research EC of Hacettepe University (2020/11-47) 3. University Hospital of Leuven, Belgium: Ethical Committee for Research of University Hospitals Leuven (S64411) 4. Hospital Universitario La Fe in Valencia, Spain: Ethical Committee of medical investigations of Hospital Universitario La Fe (2020-498-1) 5. University Hospital Bicetre Paris-Sud, Paris, France: The AP-HP Direction de la Recherche Clinique et de l'Innovation reviewed the application and concluded on 4th of February 2021 that that no approval is needed to participate to the EPIC-PCD cohort as the Jarde law that regulates clinical research in France does not apply to the study. I confirm that all necessary patient/participant consent has been obtained and the appropriate institutional forms have been archived, and that any patient/participant/sample identifiers included were not known to anyone (e.g., hospital staff, patients or participants themselves) outside the research group so cannot be used to identify individuals.

