## Supplement material for "Association between upper and lower respiratory disease among patients with primary ciliary dyskinesia: an international study"

**Table S1**: Respiratory symptoms of past 3 months reported by EPIC-PCD participants, overall and by age group (N=457)

| | **Total N (%)** | **Age 0–6 y N (%)** | **Age 7–14 y N (%)** | **Age 15–30 y N (%)** | **Age 31–50 y N (%)** | **Age >50 y N (%)** | ***p*-value[a]** |
|---|---|---|---|---|---|---|---|
| **Number of participants** | 457 (100) | 47 (100) | 149 (100) | 173 (100) | 54 (100) | 34 (100) | |
| **Nasal symptoms** | | | | | | | 0.103 |
| Daily/often | 242 (53) | 23 (49) | 77 (52) | 82 (47) | 36 (67) | 24 (71) | |
| Sometimes/rarely | 165 (36) | 16 (34) | 55 (37) | 70 (41) | 15 (28) | 9 (26) | |
| Never | 50 (11) | 8 (17) | 17 (11) | 21 (12) | 3 (5) | 1 (3) | |
| **Headache while bending down** | 50 (11) | 1 (2) | 8 (5) | 29 (17) | 7 (13) | 5 (15) | 0.014 |
| **Ear pain** | | | | | | | <0.001 |
| Daily/often | 64 (14) | 2 (4) | 13 (9) | 28 (16) | 10 (19) | 11 (32) | |
| Sometimes/rarely | 180 (39) | 15 (32) | 57 (38) | 63 (37) | 31 (57) | 14 (41) | |
| Never | 213 (47) | 30 (64) | 79 (53) | 82 (47) | 13 (24) | 9 (26) | |
| **Hearing problems** | | | | | | | <0.001 |
| Daily/often | 98 (22) | 7 (15) | 25 (17) | 25 (15) | 19 (35) | 22 (65) | |
| Sometimes/rarely | 124 (27) | 7 (15) | 45 (30) | 47 (27) | 18 (33) | 7 (20) | |
| Never | 235 (51) | 33 (70) | 79 (53) | 101 (58) | 17 (32) | 5 (15) | |
| **Shortness of breath** | | | | | | | <0.001 |
| Daily/often | 73 (16) | 1 (2) | 7 (5) | 21 (12) | 27 (50) | 17 (50) | |
| Sometimes/rarely | 206 (45) | 17 (36) | 64 (43) | 90 (52) | 20 (37) | 15 (44) | |
| Never/unknown | 178 (39) | 29 (62) | 78 (52) | 62 (36) | 7 (13) | 2 (6) | |
| **Sputum production** | | | | | | | 0.043 |
| Daily/often | 253 (55) | 18 (38) | 78 (53) | 104 (60) | 27 (50) | 26 (76) | |
| Sometimes/rarely | 137 (30) | 16 (34) | 51 (34) | 46 (27) | 18 (33) | 6 (18) | |
| Only during physiotherapy | 28 (6) | 5 (11) | 11 (7) | 10 (6) | 2 (4) | 0 (0) | |
| Never/unknown | 39 (9) | 8 (17) | 9 (6) | 13 (7) | 7 (13) | 2 (6) | |

EPIC-PCD: Ear-nose throat prospective international cohort of patients with primary ciliary dyskinesia. y: years. Symptoms are presented as N and column. [a]Chi-square test of independence.

**Table S2**: Sinonasal examination findings and prescribed treatments of EPIC-PCD participants, overall and by age group (N=457)

| | Total N (%) | Age 0–6 y N (%) | Age 7–14 y N (%) | Age 15–30 y N (%) | Age 31–50 y N (%) | Age >50 y N (%) | *p*-value[a] |
|---|---|---|---|---|---|---|---|
| **Number of participants** | 457 (100) | 47 (100) | 149 (100) | 173 (100) | 54 (100) | 34 (100) | |
| **Nasal oedema** | | | | | | | 0.025 |
| Yes | 129 (28) | 7 (15) | 40 (27) | 50 (29) | 17 (32) | 15 (44) | |
| No | 226 (50) | 22 (47) | 76 (51) | 93 (54) | 25 (46) | 10 (29) | |
| Not assessed | 102 (22) | 18 (38) | 33 (22) | 30 (17) | 12 (22) | 9 (27) | |
| **Nasal polyps** | | | | | | | <0.001 |
| Yes | 66 (14) | 2 (4) | 12 (8) | 26 (15) | 17 (31) | 9 (26) | |
| No | 338 (74) | 30 (64) | 111 (75) | 136 (79) | 36 (67) | 25 (74) | |
| Not assessed | 53 (12) | 15 (32) | 26 (17) | 11 (6) | 1 (2) | 0 (0) | |
| **Nasal discharge** | | | | | | | 0.586 |
| Yes | 345 (76) | 32 (68) | 113 (76) | 127 (73) | 44 (81) | 29 (85) | |
| No | 98 (21) | 12 (26) | 31 (21) | 42 (24) | 9 (17) | 4 (12) | |
| Not assessed | 14 (3) | 3 (6) | 5 (3) | 4 (2) | 1 (2) | 1 (3) | |
| **Type of nasal discharge[b]** | | | | | | | 0.889 |
| Serous | 111 (32) | 13 (41) | 40 (35) | 37 (29) | 11 (25) | 10 (35) | |
| Sero-mucous | 147 (43) | 12 (38) | 47 (42) | 54 (43) | 22 (50) | 12 (41) | |
| Muco-purulent | 73 (21) | 6 (19) | 21 (19) | 31 (24) | 9 (20) | 6 (21) | |
| Mixed with blood | 4 (1) | 1 (3) | 1 (1) | 1 (1) | 0 (0) | 1 (3) | |
| Not described | 10 (3) | 0 (0) | 4 (3) | 4 (3) | 2 (5) | 0 (0) | |
| **Nasal corticosteroids** | | | | | | | <0.001 |
| Yes | 101 (22) | 3 (6) | 21 (14) | 41 (24) | 24 (44) | 12 (35) | |
| No | 286 (63) | 34 (72) | 110 (74) | 112 (65) | 20 (12) | 10 (30) | |
| Not described | 70 (15) | 10 (21) | 18 (12) | 20 (12) | 10 (19) | 12 (35) | |
| **Nasal rinsing** | | | | | | | <0.001 |
| Yes | 221 (48) | 12 (25) | 76 (51) | 89 (51) | 30 (56) | 14 (41) | |
| No | 134 (29) | 21 (45) | 52 (35) | 48 (28) | 7 (13) | 6 (18) | |
| Not described | 102 (22) | 14 (30) | 21 (14) | 36 (21) | 17 (31) | 14 (41) | |
| **Inhaled corticosteroids** | | | | | | | <0.001 |
| Yes | 153 (34) | 8 (17) | 50 (34) | 66 (38) | 19 (35) | 10 (29) | |
| No | 193 (42) | 16 (34) | 60 (40) | 82 (47) | 24 (44) | 11 (32) | |
| Not described | 111 (24) | 23 (49) | 39 (26) | 25 (14) | 11 (20) | 13 (38) | |
| **Prophylactic antibiotic** | | | | | | | <0.001 |
| Yes | 141 (31) | 4 (9) | 42 (28) | 49 (28) | 28 (52) | 18 (53) | |
| No | 214 (47) | 24 (51) | 67 (45) | 100 (58) | 14 (26) | 9 (26) | |
| Not described | 102 (22) | 19 (40) | 40 (27) | 24 (14) | 12 (22) | 7 (21) | |

EPIC-PCD: Ear-nose throat prospective international cohort of patients with primary ciliary dyskinesia. y: years. Characteristics are presented as N and column %, age as median and IQR: interquartile range. [a]Chi-square test of independence. [b]Of 345 participants with nasal discharge found at examination.

**Table S3:** Results of transmission electron microscopy (TEM) of EPIC-PCD participants, overall and by age group (N=232)

| | Total N (%) | Children <18 y N (%) | Adults ≥18 y N (%) |
|---|---|---|---|
| **Number of participants** | | | |
| **Hallmark defects**[a] | | | |
| ODA and IDA-defect | 70 (30) | 50 (35) | 20 (22) |
| ODA-defect | 38 (16) | 21 (15) | 17 (19) |
| Microtubular disorganisation and IDA defect | 35 (15) | 21 (15) | 14 (16) |
| **Non-hallmark defects**[b] | | | |
| Central complex defect | 20 (9) | 4 (3) | 16 (18) |
| Other | 26 (11) | 17 (12) | 9 (10) |
| Normal ultrastructure | 43 (18) | 29 (20) | 14 (16) |

EPIC-PCD: Ear-nose throat prospective international cohort of patients with primary ciliary dyskinesia. y: years. Results presented as N and column %. ODA: outer dynein arm. IDA: inner dynein arm defect. [a]Hallmark TEM defects: ODA, ODA and IDA, microtubular disorganisation and IDA defects (based on BEAT-PCD TEM criteria by Shoemark et al). [b]Non-hallmark TEM defects included class 2 defects such as central complex defects.

**Table S4:** Association of shortness of breath and sputum production with chronic rhinosinusitis (CRS) in EPIC-PCD participants with available transmission electron microscopy (TEM) results (N=232)

| | Odds ratio | 95% confidence interval | *p*-value |
|---|---|---|---|
| | **Shortness of breath** | | |
| **CRS** | 2.49 | 1.46 – 4.24 | 0.001 |
| **TEM** | | | |
| ODA & IDA defect | | Reference | |
| ODA defect | 1.76 | 0.82–3.67 | 0.133 |
| Microtubular disorganisation and IDA defect | 1.15 | 0.56–2.37 | 0.696 |
| Non-hallmark defects | 1.31 | 0.67–2.57 | 0.424 |
| Normal ultrastructure | 0.81 | 0.41–1.61 | 0.553 |
| | **Sputum production** | | |
| **CRS** | 1.75 | 1.05–2.92 | 0.033 |
| **TEM** | | | |
| ODA & IDA defect | | Reference | |
| ODA defect | 1.62 | 0.78–3.37 | 0.198 |
| Microtubular disorganisation and IDA defect | 0.90 | 0.44–1.84 | 0.765 |
| Normal ultrastructure | 1.50 | 0.75–2.98 | 0.250 |
| Non-hallmark defects | 1.08 | 0.56–2.09 | 0.815 |

EPIC-PCD: Ear-nose throat prospective international cohort of patients with primary ciliary dyskinesia. CRS – chronic rhinosinusitis composite exposure variable consisting of 1) daily or often reported nasal symptoms and 2) examination findings of nasal discharge (sero-mucous, muco-purulent, or mixed with blood) or nasal oedema. ODA: outer dynein arm. IDA: inner dynein arm defect. Hallmark TEM defects: ODA, ODA and IDA, microtubular disorganization and IDA defects (based on BEAT-PCD TEM criteria by Shoemark et al). Normal ultrastructure referred to any non-pathologic TEM result. Non-hallmark TEM defects included class 2 defects such as central complex defects.

**Table S5:** Association of chronic rhinosinusitis with $FEV_1$ z-score among EPIC-PCD with available lung function and transmission electron microscopy (TEM) results (N=152)

| | Coefficient | 95% confidence interval | *p*-value |
|---|---|---|---|
| **CRS** | 0.18 | -1.36–1.71 | 0.820 |
| **Age** | 0.04 | -0.01–0.10 | 0.102 |
| **TEM** | | | |
| ODA & IDA defect | | Reference | |
| ODA defect | 1.38 | -0.73–3.48 | 0.198 |
| Microtubular disorganisation & IDA defect | -1.10 | -3.21–1.01 | 0.304 |
| Normal ultrastructure | 1.07 | -0.97–0.44 | 0.301 |
| Non-hallmark defects | -1.30 | -3.24–0.63 | 0.185 |

EPIC-PCD: Ear-nose throat prospective international cohort of patients with primary ciliary dyskinesia. CRS – chronic rhinosinusitis composite exposure variable consisting of daily or often reported nasal symptoms and examination findings of nasal discharge (sero-mucous, muco-purulent, or mixed with blood) or nasal oedema. ODA: outer dynein arm. IDA: inner dynein arm defect. Hallmark TEM defects: ODA, ODA and IDA, microtubular disorganisation and IDA defects (based on BEAT-PCD TEM criteria by Shoemark et al). Non-hallmark TEM defects included class 2 defects, such as central complex defects.
